# Pharmacological class interventions for benzodiazepine withdrawal discontinuation: a meta-analysis

**DOI:** 10.1101/2020.07.07.20148403

**Authors:** Dimy Fluyau, Neelambika Revadigar, Paroma Mitra, Christopher G. Pierre

**Affiliations:** Emory University School of Medicine, Board certified of Addiction medicine and Psychiatry, Emory University, School of Medicine. Brain Health.1648 Pierce Dr NE, Atlanta, GA 30307. Contact information: or.; Columbia University, Department of Psychiatry, Milstein 9 Garden North, Mail Code: 177 Fort Washington Avenue, New York NY 10032, United States, Contact.; Board certified of Psychiatry and Geriatrics, NYU, Langone Health. Department of Psychiatry.1 Park Avenue, 8th Floor New York, NY 10016., Contact. 617 -997 -3458; Grady Memorial Hospital. Emergency Department. 80 Jesse Hill Jr Dr SE, Atlanta, GA 30303. Contact. - 8210

**Keywords:** benzodiazepine withdrawal, benzodiazepine discontinuation, benzodiazepine cessation, benzodiazepine dependence

## Abstract

**Background:** Long-term benzodiazepine (BZD) use may lead to dependence, addiction, and neuropsychiatric disturbances. BZD discontinuation can cause severe withdrawal symptoms and resurgence of premorbid conditions. There are guidelines on how to stop BZD if it is necessary.

Pharmacological management is an option among several other recommendations, but its benefit remains unclear. This study investigates whether certain pharmacological classes can manage or facilitate BZD withdrawal beyond BZD itself.

**Methods:** Data collected from (1985 to 2018) in Google Scholar, Medline Ovid, Scopus, PsychInfo, ClinicalTrials.gov, Cochrane Review Database, Embase, Scopus, Pubmed, and Proquest databases: involved controlled clinical trials on drugs studied for BZD withdrawal discontinuation. Single drugs were clustered into their pharmacological class (domain). The Oxford Quality Scoring System assessed the quality of a trial. The GRADE (Grading of Recommendations, Assessment, Development, and Evaluations) was used for clinical practice recommendations. For publication bias, we visually inspected the Funnel plot. We adopted the Cochrane Risk of Bias Tool to assess biases inherent to individual trials. The standardized mean difference measured the magnitude of the benefit of a pharmacological class.

**Results:** We analyzed forty-nine controlled trials of 2815 assigned participants. Of fourteen classes, the BZD receptor antagonist class (d 0.671, CI 0.199 -1.143, p=0.005, I^2^=0),5-HT1A receptor partial agonist, and the glutamate class seemed to have the potentiality to manage BZD withdrawal discontinuation clinically. Around 61 % of the trials received an Oxford Quality score of three, 86% of the trials were granted a GRADE recommendation low. About 29 trials were at low risk of bias in general.

**Conclusions:** Even though we could not prove that the pharmacological classes of drugs we analyzed for the clinical management of BZD withdrawal discontinuation were efficacious, our investigation showed that some of these classes have the potentiality to manage BZD withdrawal discontinuation and clinically facilitate the process when it is necessary, relevant, and recommended based on established guidelines. Further investigations are warranted to support our findings.

## Introduction

The widespread use of benzodiazepines (BZDs) has been the topic of attention since the 1980s (1). A higher risk of death by suicide was observed among patients with long-term BZD use (2). There is a high prevalence of BZD use around the world. In Germany, the estimate BZD-dependent persons ranged from 128 000 to 1.6 million. Most estimates did not include many private prescriptions (3). In France, around 12.5% of patients older than eighteen were prescribed BZDs at least once during 2006(4). Australia assisted in a 21% increase of BZD use from years 2000– 2006 in the elderly with low socioeconomic status. In Thailand, 50% of the physicians prescribed BZDs for more than 25% of their patients (5). Between 2002-2014, the number of patients receiving BZDs in the United States of America increased to a greater extent than the number of patients receiving opioids. BZD use is comorbid with infections such as human immunodeficiency virus (HIV) and hepatitis C (6, 7). Medical and neuropsychiatric complications such as falls, confusion, depression, memory loss, and sleepiness are debilitating. BZD prescriptions have legal ramifications and criminal responsibilities as well as clinical and ethical concerns when one discontinues the medication against the patient’s wishes. Some authors suggested to discontinue psychotropic medications slowly as a prudent clinical and research policy (8). In most of the states in the United States of America; physicians are obligated to check the Prescription Drug Monitoring Program (PDMP) database before prescribing BZDs.

Continued BZD use can cause dependence. Attempts to discontinue the drug can lead to withdrawal discontinuation, thus the re-emergence of premorbid conditions (anxiety, insomnia, panic attacks…) and subsequently the reintroduction of the drug. The pharmacological mechanism of BZD withdrawal is complicated and unclear (9). Several experimental mechanistic approaches of BZD discontinuation were the subject of research; most of the theory targeted BZD withdrawal. It is suggested that upregulation of the gamma-aminobutyric acid (GABA-A) receptor binding complex is one of the molecular mechanisms for BZD discontinuation (10). However, other hypotheses have been the subject of many publications. The alterations in GABA-A receptor subunit expression suggested that chronic BZD exposure led to impaired sensitivity by down-regulating the response of a drug to the GABA-A receptors, mainly GABA-A receptors that contain an α1, α2, α3, or α5 subunit (11, 12). Other neurobiological components are also involved in BZD discontinuation such as glutamate, transcriptional and neurotrophic factors (GABARAP, BIG2, PRIP, gephyrin, and radixin), serotonin, dopamine, acetylcholine receptor systems and neurosteroids complex (13).

There are several medications (buspirone, lithium, atenolol, carbamazepine, flumazenil) investigated for the management of BZD withdrawal discontinuation. Carbamazepine, for example, was suggested in two systematic reviews (years: 2006 and 2018) (14,15) to have the potentiality to control withdrawal discontinuation. A meta-analysis suggested that flumazenil successfully facilitated BZD discontinuation (year:2006) (16). There is still inconclusive evidence that these treatments are effective. The previous investigations focused on a single pharmacological agent for BZD discontinuation. We attempted another approach by clustering single drugs into their pharmacological classes (glutamate, BZD receptor antagonist…) according to the Neuroscience-based Nomenclature (NbN).

Several debates and publications reflected that the World Health Organization (WHO) psychopharmacological nomenclature does not reflect the contemporary developments and knowledge relevant to brain disorders. The WHO’s nomenclature can choose an “antipsychotic” to treat both depression and schizophrenia. Such intervention can confuse the patients and compromise their adherence to treatment (17,18). In contrast, the NbN identifies the pharmacological drug target, mechanism, and family that reflect the primary neurotransmitter, neurobiological activities (neurotransmitter, brain circuits, and physiological effects), and details clinical observations (the drug efficacy and side effects) (18). Referring to the NbN; Nutt and colleague suggested that a” better understanding of pharmacology can benefit translational neuroscience and the discovery of new treatments for brain disorders” (19).

This study investigates whether certain pharmacological classes can manage or facilitate BZD withdrawal beyond BZD itself.

## Methods

Figure 1 outlines the search process. Ethical approval was not required because the study was a secondary analysis of anonymized data that were already published. The meta-analysis was not registered in systematic review databases.

**Figure 1.**
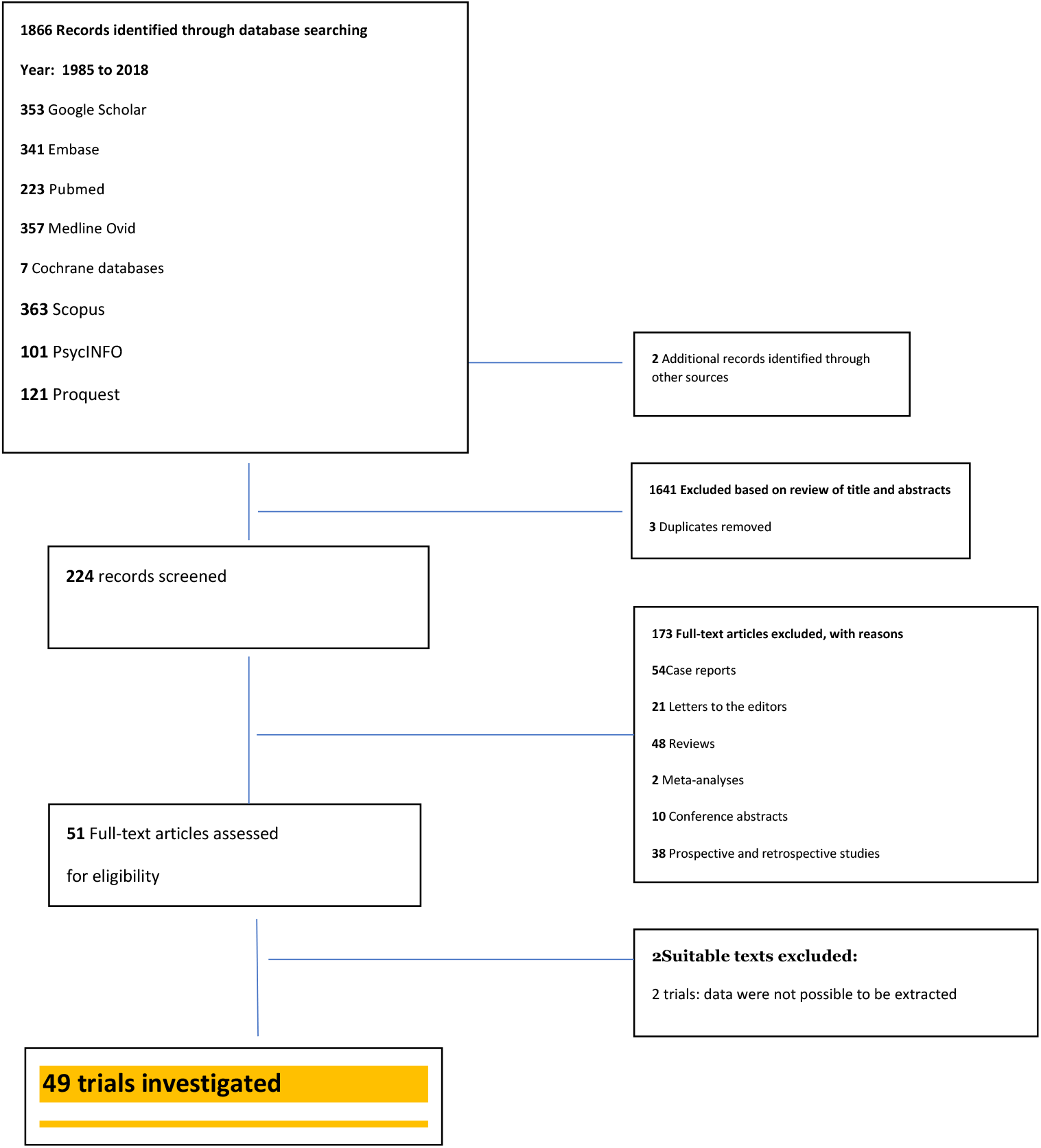
Outline of the search process; included articles as well as excluded articles with reasons for exclusion.

### Included

We selected published controlled clinical trials that compared an active drug with a placebo or with another active drug or a control(non-placebo) in participants with a history of long-term BZD use or BZD-dependent who are undergoing BZD discontinuation.

### Excluded

We excluded cross-sectional, longitudinal studies, and uncontrolled trials.

### Types of participants

The participants were approximately 18 years of age or older, male or female, on a BZD for medical or neuropsychiatric reasons.

### Types of interventions

Controlled trials evaluating any active drug to facilitate BZD withdrawal discontinuation.

### Control intervention

Controlled trials comparing an active drug with a placebo or with another drug or a control(non-placebo) to facilitate BZD discontinuation.

### Types of outcome measures

Investigate the effectiveness (reduction of anxiety or depressive symptoms, improve sleep, decrease irritability or aggression) of drugs (clustered into pharmacological classes) studied for the clinical management of BZD withdrawal symptoms during the discontinuation period.

### Types of settings

The investigations were conducted in outpatients, hospital, multicenter, psychopharmacology research unit, research centers, and specialized clinics (methadone clinic, for example).

### Electronic searches

We integrated the Boolean logic strategy by free-texting: “benzodiazepine withdrawal, benzodiazepine withdrawal discontinuation, benzodiazepine discontinuation, benzodiazepine discontinuation trial, and benzodiazepine withdrawal trial” in Google Scholar, Medline Ovid, Scopus, PsychInfo, ClinicalTrials.gov, Cochrane Review Database, Embase, Scopus, Pubmed, and Proquest by customizing articles by dates of publications (from 1985 to 2018). A search in the Grey Literature was also undertaken using the above strategy at Open Grey.

### Searching other resources

We searched for other sources of trials. We were able to find two additional copies of publications that we could not find via the electronic search. We could not find unpublished trials despite our effort in contacting two drug manufacturers.

### Data collection and analysis

#### Selection of studies

Two authors (DF and PM) independently searched for publications, screened abstracts, full articles, and extracted data using the AbstractionForm (20) on the intervention components, methods of blinding and randomization, outcomes of interest, study design, sample size, and age. The two authors also scored the quality of the trials by using the Oxford Quality Scoring System (21) (≤ 2: low range of quality score, ≥ 3: high range of quality score, and 5: highest score) and granted a GRADE recommendation level for each trial (22) (Refer to table 1). Two other authors (CP and NR) settled disagreements between DF and PM.

**Table 1.**
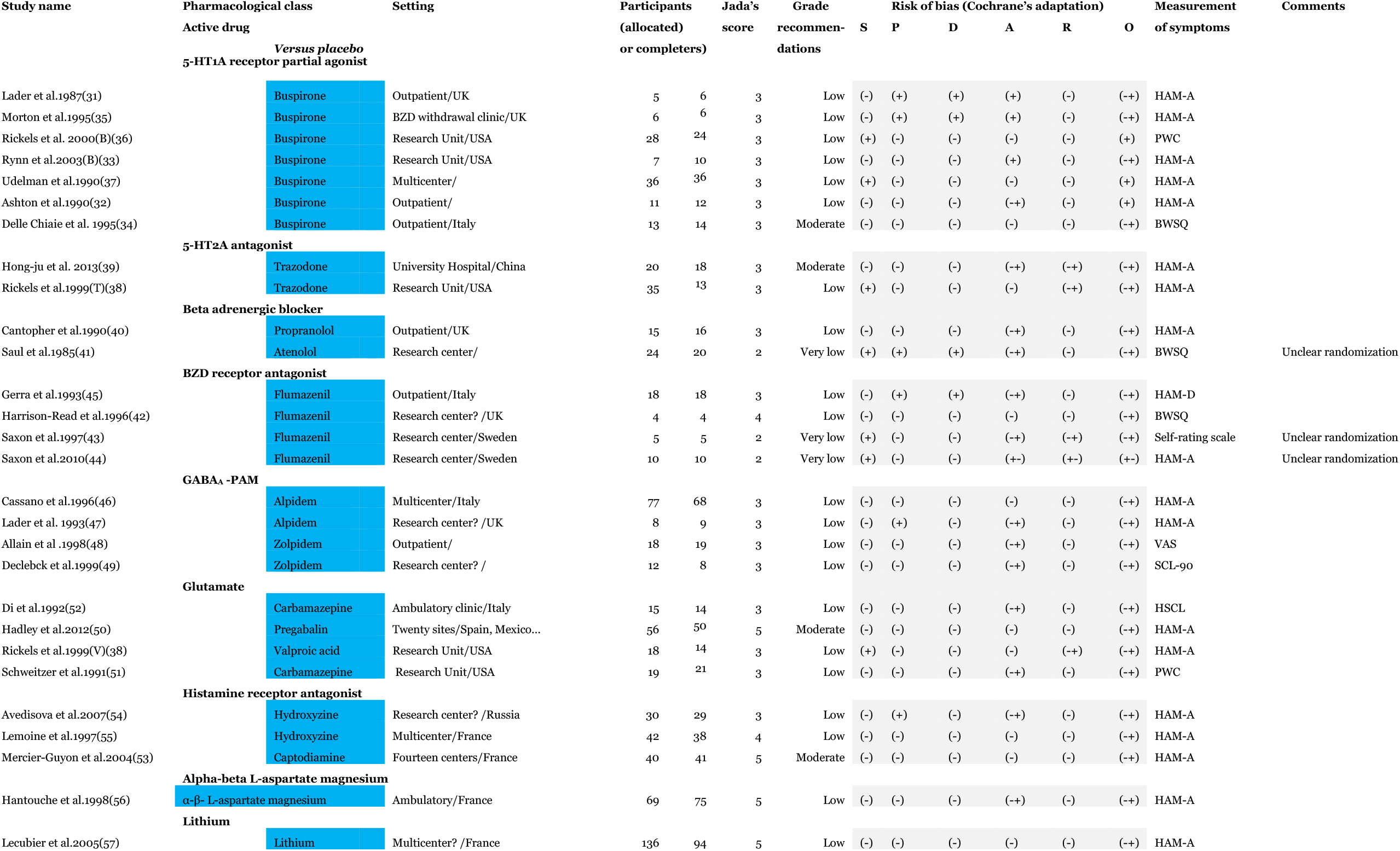
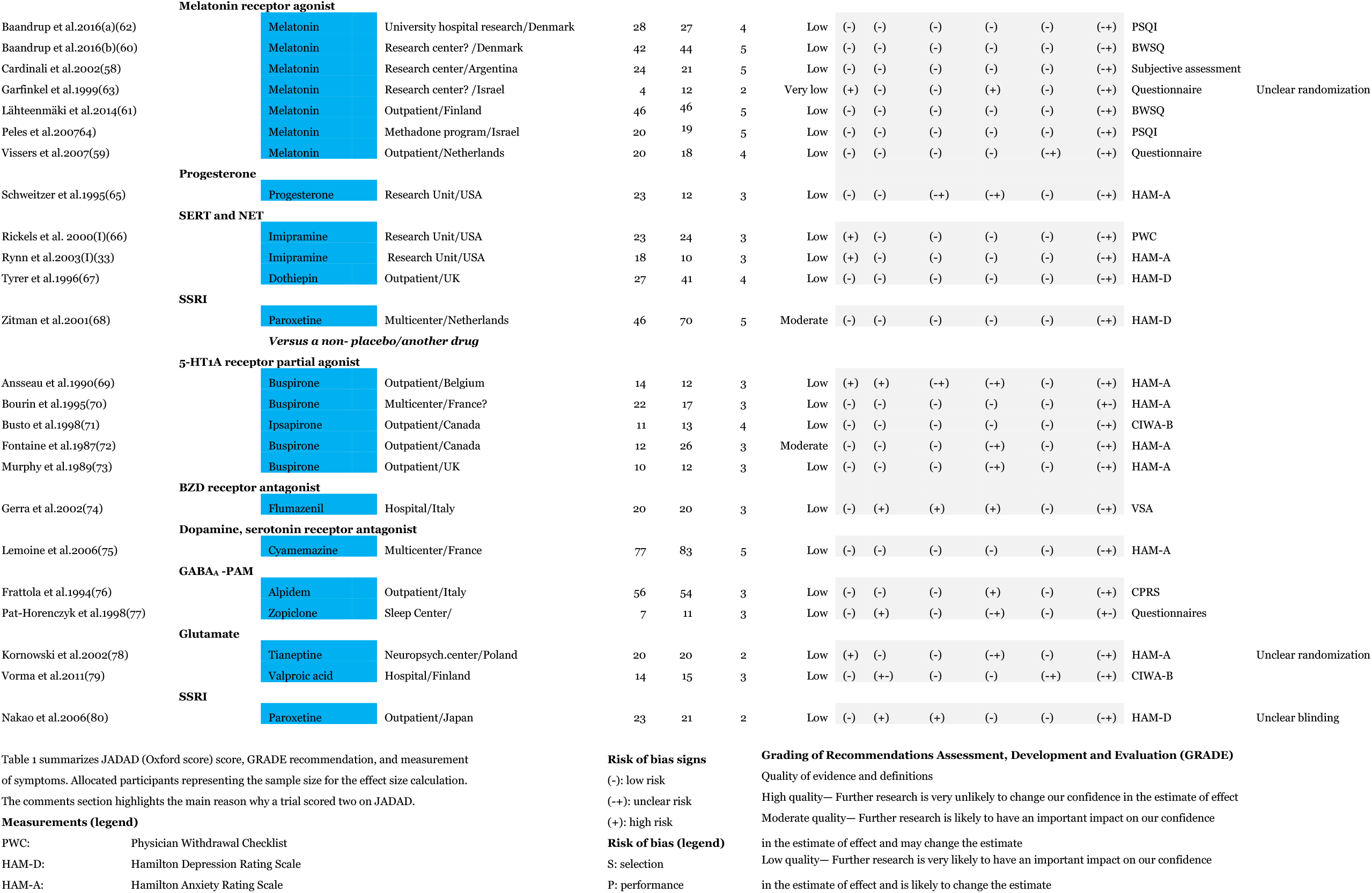
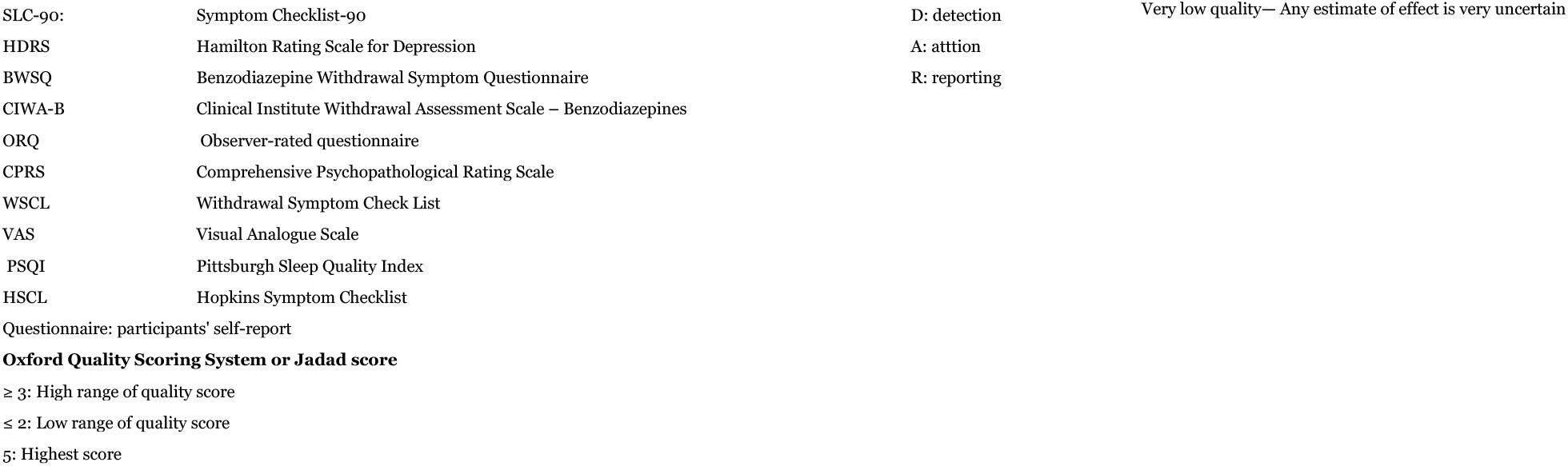

### Measures of treatment effect

We calculated, recalculated, or extracted the mean and standard deviation score of a trial. If the mean and standard deviation was plotted in a graph, we used the WebPlotDigitizer (23) to extract data so we could convert them later to the mean and standard deviation. For the group effect, we opted for the random effect model (24) because of sampling variability and difference in treatment effect of individual trials. For the measure of treatment, we adopted the standardized mean difference between the two groups (Cohen’s d). Cohen’s d =0.2 was interpreted as a small benefit or advantage, 0.5 was medium, and 0.8 was large.

### Unit of analysis issues

None.

### Dealing with missing data

We contacted two authors for missing data; however, only one author replied and said that the requested data could not be found.

### Assessment of heterogeneity

We used I^2^ to quantify the dispersion of effect size. An I^2^ of 25% was low, 50% was moderate, and 75% was high (20).

### Assessment of biases

For **publication bias**, we visually inspected the funnel plot for symmetry or asymmetry and for the distribution of small studies (smaller sample size) in comparison to more extensive studies. We drew some suggestions on the possibility that publication bias might show asymmetry in the funnel plot, and that smaller studies and positive studies had more chance to be published. The funnel plot might not be an accurate representation of publication bias; its interpretation should be considered inconclusive. We adopted the Cochrane risk of bias tool (25) to assess **selection, detection, attrition**, and **performance biases** of individual trials (Table 1).

### Data synthesis

The data we analyzed were combinable based on both outcome measures and neuropharmacological mechanism of action. We conducted the statistical analyses in the Comprehensive Meta-Analysis software, version 3(26) (14 North Dean Street, Englewood, NJ 07631 USA, and OpenMEE (27) (British Ecological Society, Charles Darwin House, 12 Roger St, London, WC1N 2JU, UK) with the latest version for Windows 10.

### Subgroup analysis and investigation of heterogeneity

We clustered single drugs into their respective pharmacological classes and compared them with placebo or with another active drug or a control(non-placebo). We hypothesized that such methodology could lessen heterogeneity and make the data more analyzable and clinically relevant.

### Sensitivity analysis

We performed a sensitivity analysis for subgroups with a minimum of four studies whenever it was possible.

## Results

### Trials included and their characteristics

Our search (in line with the Preferred Reporting Items for Systematic Reviews and Meta-Analyses (PRISMA) (28) indicated 1866 published studies from 1985 to 2018 and two other studies additional identified through other sources. After removal of 1641 studies based on titles, abstracts, and duplicates, the literature yielded 224 potentially relevant trials. Subsequently, 173 trials of the 224 were excluded for various reasons (Fig. 1). In total, forty-nine controlled clinical trials were selected and included in this meta-analysis after two suitable one (29,30) were removed because we could not extract enough data, in conformity to the measures of treatment effect that we opted to perform statistical analysis. The trials involved 2724 assigned participants (allocated or completers):1384 participants in the active medication group and 1340 in the placebo group, another group or a control group(non-placebo) (Table 1). Eleven trials studied buspirone, six trials studied melatonin, and five trials studied flumazenil. The mean sample size for the medications was 26.61, with a margin error:95% CI, 1.960σ_x□_, 26.6154±6.367 (±23.92%). The mean sample size for the placebo or another active drug(non-placebo) was 25.76, with a margin error: 95% CI, 1.960σ_x□_, 25.7692 ±5.669(±22.00%).

### Study quality

On average, 61 % of the trials received an Oxford Quality score of three; 20% of them received a score of five; 11% received a score of four, and less than 12% of them received a score of two. Around 86% of the trials were granted a GRADE recommendation low (Table 1). We judged that 21 % of the trials were at high risk for selection bias due to inadequate generation of a randomized sequence trial, and 19% of them were at high risk of performance bias due to insufficient description of blinding procedures for participants and personnel. Around 12% of the trials were at high risk for detection bias due to insufficient information about the knowledge of the allocated interventions by outcome assessors. In 41% of trials, attrition bias was unclear, though we judged that around 12% of the trials had a low risk of attrition bias. Overall, 86% of the trials had a low risk of reporting bias, and about 29 trials were at low risk of bias in general (Table 1).

### Meta-analysis involving placebo as a comparator (Forest plot 1)

#### 1. Management of anxiety

##### 1.1. HT1A receptor partial agonist

The 5-HT1A receptor partial agonist class involved seven trials. In two trials, buspirone did not alleviate BZD-withdrawal symptoms, and there was a high dropout compared with placebo (31, 32). During BZD discontinuation buspirone showed no difference to placebo at attenuating anxiety symptoms severity (33). In other trials, buspirone showed no BZD-withdrawal symptom in participants off lorazepam (34), and the drug lowered anxiety level (35). Data collected for buspirone showed a modest reduction in symptoms of anxiety and depression during BZD-tapered (36). Anxiety was significantly reduced in the buspirone group at the beginning of alprazolam withdrawal and endpoint (37). The class’s effect size was very small (d 0.086). The result was not statistically significant(p=0.711). Heterogeneity was moderate: I^2^= 65.99%. The margin of error of the sample size for the five studies was estimated to be around 32.8 ±21.694(95%, 1.960σx□). Leaving out two studies with sample size ≤ 32 increased the effect size (d 0.119) but remained statistically non-significant(p=0.776). The effect size was large (d 3.000) when studies were adjusted based on a high dropout rate. The heterogeneity lessened (I^2^=53.014%).

##### 1.2. HT2A antagonist

The 5-HT2A antagonist involved two trials. Trazodone did not reduce withdrawal severity when compared with placebo. Participants complained of sedation and dry mouth (38). Another trial reported that trazodone lower BZD-withdrawal symptoms based on the interpretation of scores of the Withdrawal Symptoms Checklist. Also, no adverse effect was found (39). There was no apparent benefit of 5-HT2A antagonist (d 0.068). There was no heterogeneity: I^2^=0%.

##### 1.3. Beta-adrenergic blocker

This class included two trials. Propranolol(non-cardioselective) did not reduce BZD withdrawal symptoms in BZD-dependent patients over ten weeks (40). Atenolol(cardioselective) ameliorated both the affective and somatic symptoms of anxiety (41). The b-blockers showed a small effect size benefit to lessen somatic symptoms during BZD withdrawal (d 0.107), with a high degree of imprecision (95% CI −0.347-0.562,p= 0.644).

##### 1.4. BZD receptor antagonist

In an interrupted trial in which flumazenil precipitated panic and dysphoria, it was suggested that the drug could potentially benefit patients dependent on BZDs (42). Other trials found that flumazenil could treat BZD withdrawal in BZD-dependent patients (43) and reduce aggression and hostility during BZD discontinuation (44). Mean withdrawal symptoms were not elevated in comparison to placebo in chronic use of a high dose of flunitrazepam or lormetazepam (45). In four trials, BZD receptor antagonist could potentially abate BZD withdrawal symptoms during BZD discontinuation period (d 0.671, CI 0.199 -1.143). The overall benefit of flumazenil was statistically significant(p=0.005). There was no heterogeneity: I^2^=0%. The funnel plot appeared to be symmetric, suggesting no publication bias. The observed outcome yielded a significant p-value =0.0033, and the target was p <0.05(Funnel plot 1).

##### 1.5. GABA_A_ receptor positive allosteric modulator

The GABAA -PAM included four trials. Alpidem (46,47), and zolpidem (48,49) clustered into their pharmacology class showed limited efficacy in alleviating withdrawal symptoms (e.g., withdrawal insomnia) during BZD discontinuation, but the effect size was small, and the result was not statistically significant (d= 0.108, p=0.479). Heterogeneity was small: I^2^= 8.261%. Analysis of high dropout due to poor tolerance and adverse events remained non-statistically significant(p=0.912).

##### 1.6. Glutamate: carbamazepine pregabalin, and valproic acid

The glutamate class included four trials. One trial found that pregabalin 300–600mg/day could help in discontinuing BZD in BZD-dependent patients (50). In a joint trial with trazodone, valproic acid did not lessen withdrawal symptoms in patients of long-term BZD use (38). On the contrary, carbamazepine lowered the incidence of withdrawal symptoms in patients who used BZD for an extended period (51,52). The class could lessen withdrawal symptoms during BZD discontinuation (d 0.344, CI 0.065-0.623,p= 0.016). Heterogeneity was null: I^2^= 0%. There was no indication of publication bias on the funnel plot (Funnel plot 2). The drugs used in the four trials were well-tolerated, and the trials were apparently of high quality, except that the sample size (allocated participants) of three studies (38,50,52) was altogether below 40(95%,1.960σx□: 33.6667 ±5.254), leaving one study with an estimated sample size above 50(95%,1.960σx□: 53 ±4.158) (51). Adjusting studies with sample size below 40, the benefit of glutamate for BZD withdrawal became non-statistically significant(p=0.063).

##### 1.7. Histamine receptor antagonist

The histamine domain included three trials. Captodiamine addressed the prevention of emergence of a BZD withdrawal during discontinuation (53). Hydroxyzine was investigated as a substitute for BZD withdrawal, and as a method to wean patients off lorazepam (54,55).

Although histamine’s effect size showed that the class could aid in managing BZD withdrawal symptoms (d 0.329), there was a broad level of imprecision where the actual effect lies (95% CI −0.188-0.0.845).

##### 1.8. alpha-beta L-aspartate magnesium

We analyzed one trial that evaluated the efficacy of alpha-beta L-Aspartate Magnesium (Asp Mg) for BZD withdrawal discontinuation (56). L-aspartate Mg’s ability to reduce the intensity of BZD withdrawal during the cessation period was not demonstrated (d 0.070).

##### 1.9. Lithium

In one trial, gluconate lithium (57) did not facilitate benzodiazepine withdrawal discontinuation among patient treated with less or equal 10 mg of benzodiazepine (d −0.047), and there was more placebo effect (60% success rate).

##### 1.10. Serotonin, norepinephrine reuptake inhibitor (SERT and NET) (dothiepin and imipramine)

The SERT and NET class included three trials (66,33,67). Overall, the benefit of the class to alleviate symptoms of depression or anxiety during BZD discontinuation was small (d 0.234). Heterogeneity was moderate I^2^=51.063.

#### 2. Management of sleep

##### 2.1. Melatonin receptor agonist

This domain included seven trials. Trials analyzed found no modification of sleep or wakefulness by melatonin after benzodiazepine withdrawal (58), no conclusive evidence that the drug could facilitate BZD discontinuation in patients with insomnia (59), no withdrawal benefit, neither could facilitate BZD discontinuation (60,61). Three trials found that melatonin could improve and maintain sleep quality during BDZ withdrawal (62,63,64). Overall melatonin showed no sleep benefit during BZD withdrawal discontinuation (d −0.059).Heterogeneity was moderate: I^2^= 57.482%.

##### 2.2. Progesterone

In one trial, micronized oral progesterone doses up to 3600 mg/day did not facilitate BZ discontinuation in BZD-dependent participants (65) (d −0.361).

#### 3. Management of depressive symptoms

##### 3.1. Selective serotonin reuptake inhibitors (SSRI) (paroxetine)

One trial evaluated the long-term outcome of an SSRI (paroxetine) after BZD withdrawal (68). Analysis of paroxetine on participants tapered off diazepam showed a small effect size compared with placebo on the Hamilton Depression Rating Scale (HAM-D) (d 0.087). This result was not statistically significant(p=0.647). However, another scale such as the State-Trait Anxiety Inventory showed a large effect size (STAI) (d 1.014), and a p-value equals to zero(p=0).

### Meta-analysis involving another drug as comparator (Forest plot 2)

#### 1. Management of symptoms of anxiety

##### 1.1. 5-HT1A receptor partial agonist (buspirone, ipsapirone)

The 5-HT1A receptor partial agonist was investigated in five comparative trials involving an active drug vs. another drug, especially another BZD. Participants who underwent 2-week withdrawal on either buspirone or oxazepam did not show a significant difference in the Hamilton anxiety scale or Hamilton Depression Rating Scale (69). No significant difference between was found buspirone vs. lorazepam in term of withdrawal discontinuation symptoms (Day 63 and Day 70) (70) and compared with lorazepam ipsapirone had fewer symptoms for withdrawal discontinuation (71). Diazepam’s participants reported more withdrawal symptoms than buspirone’s participants (72,73). Participants allotted to 5-HT1A receptor partial agonist exhibited fewer withdrawal symptoms discontinuation (d 0.369), but statistical significance was not reached (p= 0.126). Heterogeneity was low: I^2^=48.768%. Leaving out studies with a higher dropout rate did not statistically make a difference (p= 0.236). In that case, the heterogeneity was moderate: I^2^=61.558%.

##### 1.2. BZD receptor antagonist(flumazenil)

A comparison trial between oxazepam and flumazenil (74) suggested that intravenous flumazenil could reduce withdrawal symptoms such as feeling tired, tense, hungry, body aches, and heart pounding. The finding could not be substantiated based on the data we analyzed. The effect size was small (d 0.138).

##### 1.3. Dopamine, serotonin receptor antagonist (5-HT2AR, HT2CR and D2R) (cyamemazine)

In a comparative efficacy trial, cyamemazine was able to control rebound anxiety symptoms comparable to bromazepam, though the cyamemazine group had a higher dropout rate (75). Cyamemazine did not appear to be efficacious in facilitating BZD discontinuation based on the data we analyzed (d 0.044). Heterogeneity was not assessed due to a low number of trials.

##### 1.4. GABA_A_ -PAM (alpidem, zopiclone)

Alpidem produced fewer withdrawal discontinuation symptoms than alprazolam and was also better tolerated than lorazepam (76). Zopiclone was suggested to facilitate the gradual withdrawal from long-term use of long-acting BZDs. It was found that withdrawal symptoms were milder among zopiclone’s participants compared with flunitrazepam’s participants (77). Statistical significance was not reached; the GABAA -PAM’s effect size was (d 0.319). Heterogeneity was low: I^2^=23.451%.

##### 1.5. Glutamate (tianeptine, valproic acid)

In one double-blind comparison trial, tianeptine was found to be as effective and safe as carbamazepine to treat BZD withdrawal in dependent patients (78). Valproic acid could potentially reduce withdrawal symptoms in opioid-dependent patients on BZDs (79). For a total of two trials, the glutamate class potentiality at reducing BZD withdrawal symptoms was not demonstrated (d 0.149). Though, there was no heterogeneity: I^2^= 0.

#### 2. Management of depressive symptoms

##### 2.1. SSRI (paroxetine)

In one trial, paroxetine could facilitate BZD withdrawal in SSRI (+) (with paroxetine) compared with no SSRI (–) (no paroxetine) in non-depressive participants of chronic use of BZD (80). Although the effect size was medium (d 0.555), there was a large degree of imprecision (95% CI −0.048-1.157).

## Discussion

Although, we could not prove that some pharmacological classes of drugs studied for the clinical management of BZD withdrawal discontinuation were efficacious, three pharmacological classes: BZD receptor antagonist, 5-HT1A receptor partial agonist, and glutamate may be able to facilitate BZD withdrawal discontinuation management. Both BZD receptor antagonist and glutamate fared better than placebo in the primary analysis. The 5-HT1A receptor partial agonist fared better than placebo in the secondary analysis with a large effect size. For the three classes, statistical significance was not reached in the sensitivity analysis. This meta-analysis may agree and complement two previous systematic reviews (years: 2006-2018) (14,15) by the Cochrane group and one meta-analysis (year: 2006) (16) in which carbamazepine, pregabalin, and flumazenil showed positive results for BZD withdrawal. Also, lithium and melatonin (year: 2018) did not show any benefit regarding BZD withdrawal discontinuation management (14). In our meta-analysis, lithium and melatonin’s effect size were negative in the primary analysis. The study has several limitations. L-aspartate of magnesium was analyzed as if it belongs to a pharmacological class. L-aspartate of magnesium as a class by itself is hypothetical.

Carbamazepine and pregabalin may bind with different receptors resulting in different clinical benefits despite studies suggested that both drugs ‘mechanism of action may be related to the NMDA subtype of glutamate receptors (81,82). For example, carbamazepine binds to inactivated Na+ channels and reduces Ca2+and Na+ flux across the neuronal membrane (83), and pregabalin favors a selective inhibitory effect on voltage-gated calcium channels containing the α2δ-1 subunit (84). Valproic acid’s mechanism action is yet to be determined. Valproic acid causes blockade of voltage-gated sodium channels, but some studies link valproic mechanism’s action mostly to the increase of gamma-aminobutyric acid (GABA) (85). Administration of chronic lithium or carbamazepine was suggested to downregulate brain arachidonic acid signaling via NMDA receptors and may contribute to their mood stabilizer’s activity (86). Glutamatergic signaling via NMDA receptors is pathologically upregulated in bipolar disorder; valproic acid may dampen the upregulated NMDA involving arachidonic acid (86). Thus carbamazepine, pregabalin, and lithium could have been clustered into the same pharmacological class. One cannot assume that carbamazepine, valproic acid or pregabalin’s potentiality at managing BZD withdrawal is solely related to the NMDA subtype of glutamate receptor. Tianeptine’s classification as glutamate is another limitation. The drug has been suggested to be both an SSRI and SNRI. It was suggested that tianeptine does not inhibit the uptake of serotonin or noradrenaline in the central nervous system (87). Tianeptine’s effect on the glutamatergic may represent the most proximal target for the drug antidepressant’s efficacy (88).

The data we analyzed favored flumazenil. Flumazenil can itself cause severe withdrawal symptoms in long-term benzodiazepine users and seizures, especially among patients who overdose on BZD. Many practitioners remain very cautious when using flumazenil; though a study suggested that slow titration may reduce its adverse effects (89). Among the glutamate class, carbamazepine can cause leucopenia and thrombocytopenia, and pregabalin has the potentiality for abuse. Long-term use of zolpidem and zopiclone increases the risk of addiction, tolerance, dependence, and withdrawal. Thus, regardless of the outcome; zolpidem and zopiclone may have limited clinical indications for BZD withdrawal discontinuation. Another limitation is that most of the trials focused on BZD discontinuation or BZD-substitution, but some trials focused on BZD-tapered in which participants were followed-up over time.

The tool we utilized to assess the quality of the trials or to cluster medications into class also has limitations. The Oxford Quality Scoring System (Jadad scale) was criticized for possessing low interrater reliability and for being too mechanistic. The GRADE was criticized for being too subjective. The Neuroscience-based Nomenclature (NbN) is still updating, and it has not been generally adopted yet worldwide. The data extracted to calculate the effect size came from different scales that the authors utilized to report the benefit of a drug for the management of BZD discontinuation. In some cases, the authors utilized a non-standardized scale or their questionnaire. A sensitivity analysis involving a unified scale could be warranted and could have changed the results.

Stopping BZD as much as using it are both challenging. Medical and neuropsychiatric complications of BZD use, abuse, and dependence can be harmful to some patients. The British Association for Psychopharmacology (BAP) produced guidance on the risks and benefits of BZD use (90). The BAP recommended prescribing BZD for a short time. The BAP acknowledged that BZD dependence is a significant risk in some patients receiving treatment for longer than one month. Clinical judgment can guide whether alternatives (taper, psychological or pharmacological treatment) be more suitable, for each patient, and each proposed medication (90). If there is no history of drug dependence, the decision to continue BZD treatment may be more reasonable than the alternatives. Salzman and colleagues suggested that BZD prescribed for long-term use should be regularly re-evaluated, tapered and ceased when proper to decrease drug-related harm and lessen the impact of medication burden on quality of life in the elderly (91). From a clinical perspective, Soyka suggested that BZD withdrawal not need to be tried in every case, and it should be tailored to patient motivation, severe psychopathological symptoms such as depression (92). Interest should also target elderly persons. Analyzing data of a nationwide cohort study in Denmark 2000–2010, Tjagvad and colleagues found that patients with opioid use disorder were prescribed more BZDs than any other substance use disorder (93). Such data points out the imminent risk of overdose and respiratory arrest. In some parts of the United States of America, BZD prescription incurs legal consequences. BZD discontinuation seems appropriate if it is clinically warranted. A systematic review suggested that a pharmacological approach that aimed substitution to withdraw BZD with or without psychological aid had the highest success rates (94). Adequate pharmacotherapy and more evidence-based strategies are necessary during and after BZD discontinuation (92). In high dose, BZD-dependent patients’ relapse was prevalent among the one who tried to discontinue the drug. These patients perceived BZD discontinuation to be difficult, unpredictable, and complicated (95). For this category, medications that can facilitate the process may be warranted.

We present a set of statistical data as an attempt to aid the interpretation and clarification of an unclear topic by analyzing the clinical benefit of some pharmacological classes for BZD withdrawal discontinuation management. We suggest this pharmacologically driven approach that reflects the neurotransmitters and mechanism of drugs is clinically relevant and more practical. We hope our analysis can assist practitioners in making an informed decision with their patients when it is necessary to discontinue BZD and when established guidelines recommend it. This meta-analysis expands the arsenal of knowledge and evidence of the efficacy of medications for BZD withdrawal discontinuation. We believe it can illuminate the need for further researches for the benefit of the patients and make BZD discontinuation less stressful for practitioners.

## Data Availability

Data are available upon request.

## Acknowledgement

We thank Dr. Challyn Malone for the language help.

---

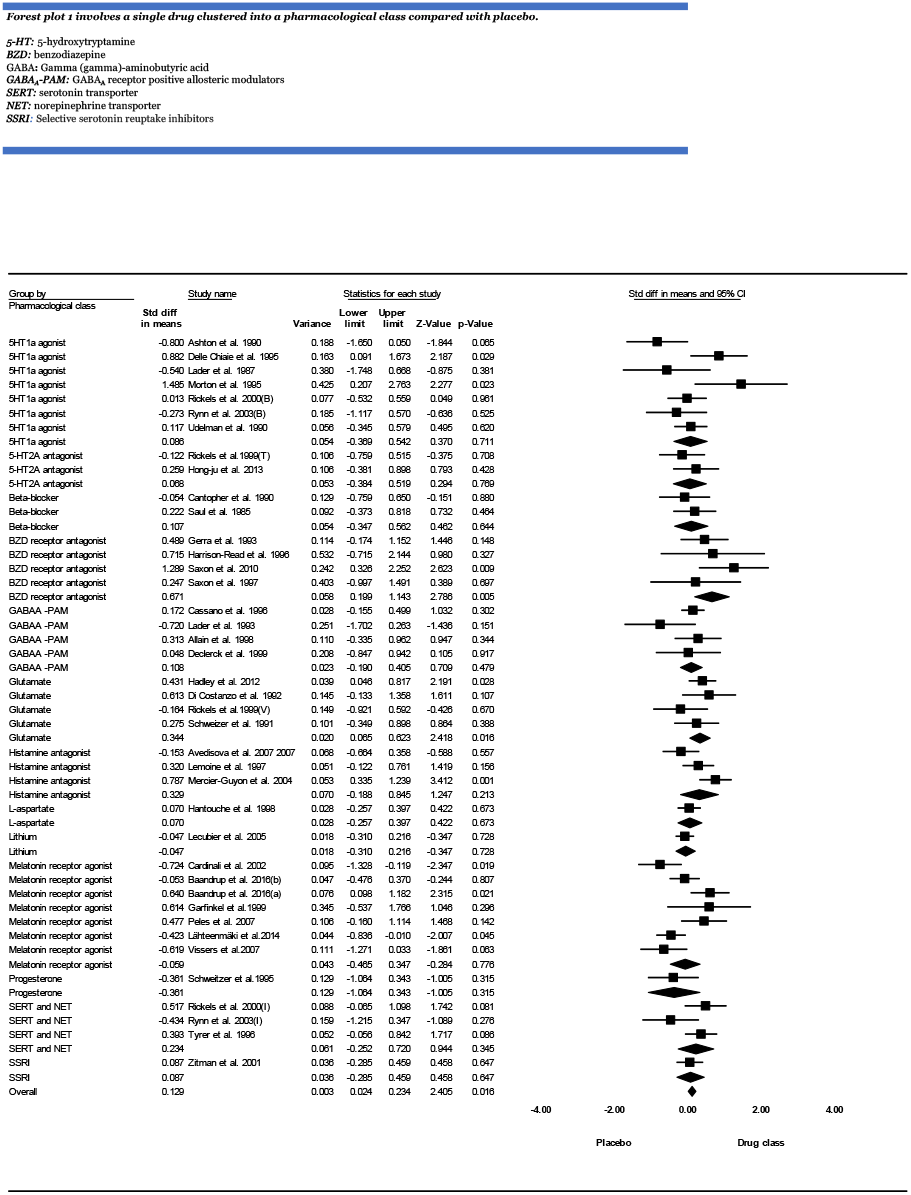

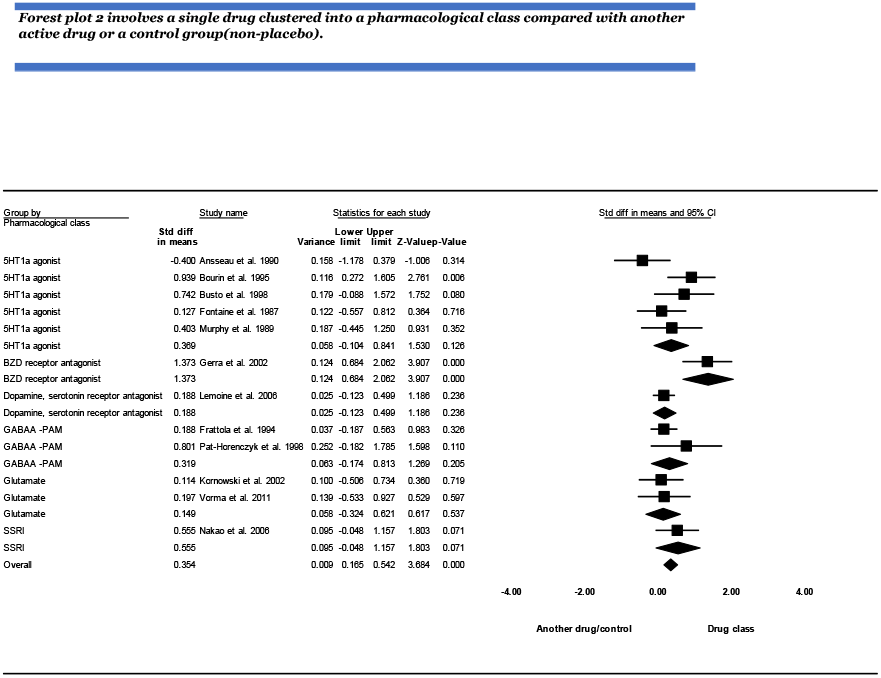

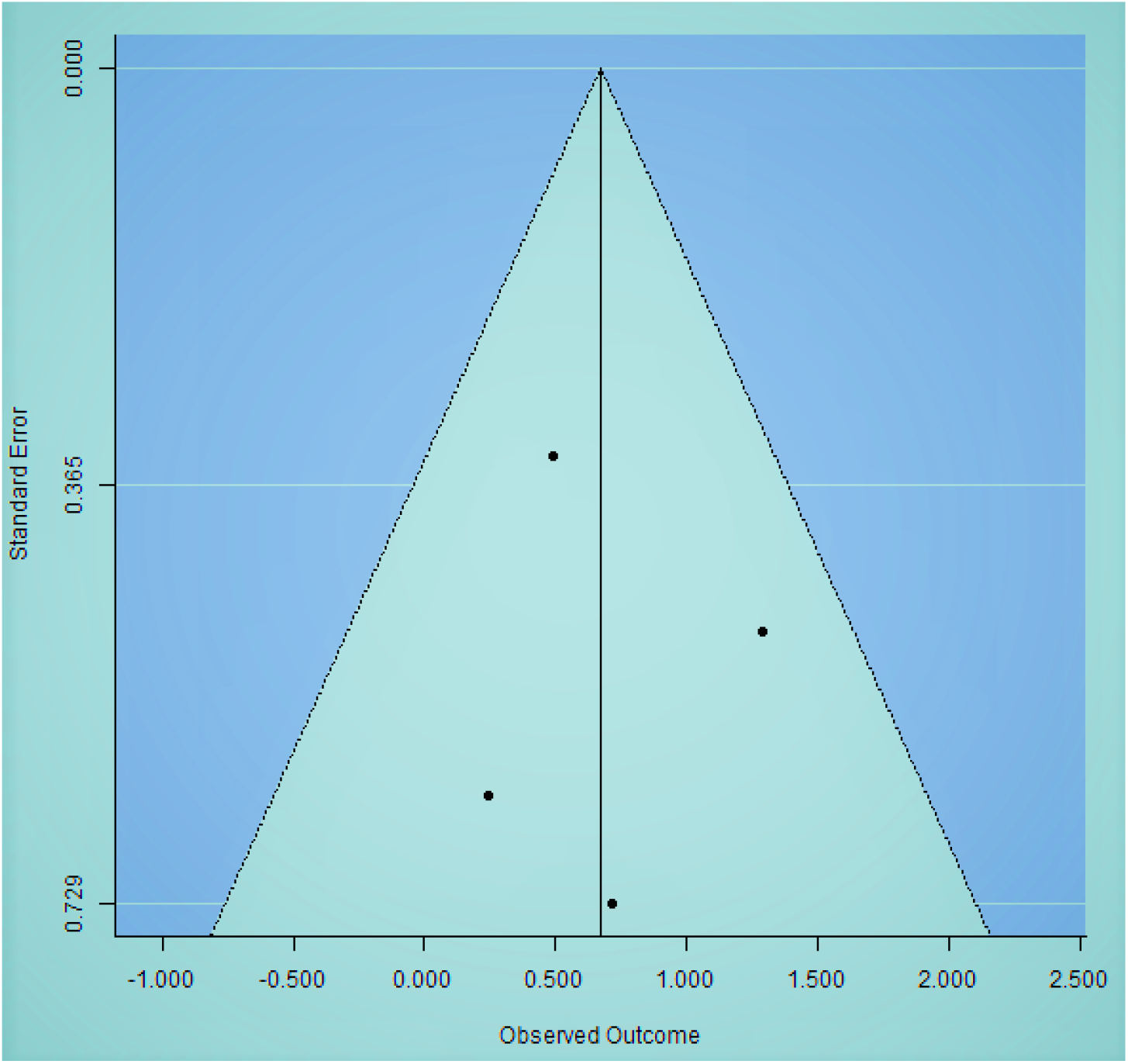

**Funnel plot 1: BZD receptor antagonist versus placebo**

**Fail-safe N: 7**

**Observed significance level:0**.**0033**

**Target significance level: 0**.**05**

The funnel plot appears to be symmetric. The observed outcome yields a significant p-value. It seems that two small studies lie below the cut off - of 0.365.

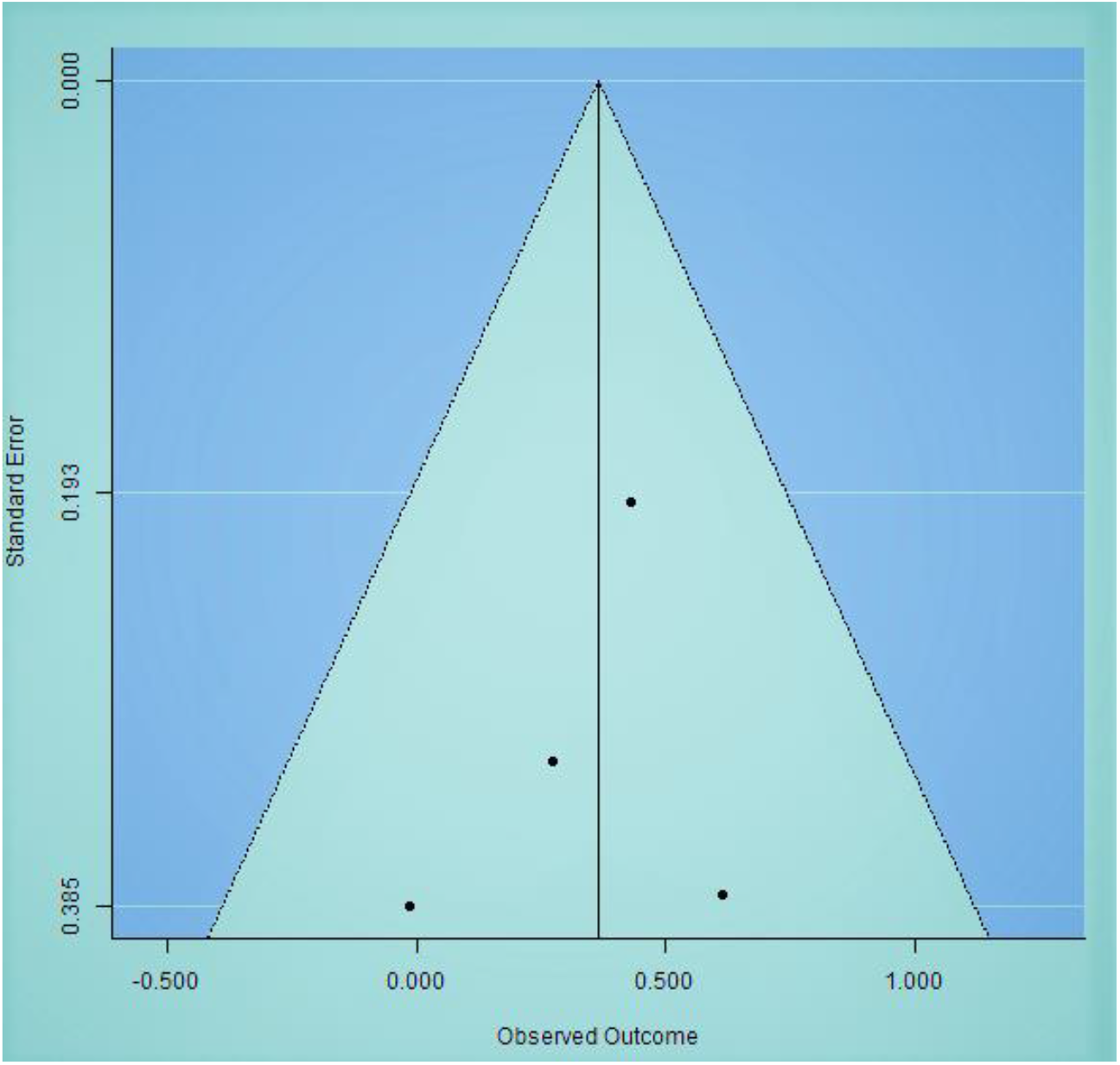

**Funnel plot 2: glutamate versus placebo**

**Fail-safe N: 4**

**Observed significance level:0**.**0103**

**Target significance level: 0**.**05**

The funnel plot appears to be symmetric. The observed outcome yields a significant p-value. It seems that three studies lie below the cut off –of 0.193 are small studies. It is more likely that the smaller studies tend to report a higher effect.

